# Prevalence of Comorbidities among United States Adults with asthma and Their Association with Asthma Severity

**DOI:** 10.1101/2023.08.27.23294694

**Authors:** Chukwuemeka E. Ogbu, Pallab Sarker, Chisa O. Oparanma, Stella C. Ogbu, Ioannis Stouras, Ebube Eze, Chinaenye Ndugba, Otobo I. Ujah, Russell S. Kirby

**Affiliations:** College of Public Health, University of South Florida, Tampa, FL 33612, USA; Department of Medicine, Sher E-Bangla Medical College, Barisal, Bangladesh; Department of Medicine, Kharkiv National Medical University, 61022 Kharkiv, Ukraine; Department of Biomedical Science, School of Medicine, Tulane University, New Orleans, LA 70112, USA; Department of Medicine, National and Kapodistrian University of Athens, Greece; Department of Internal Medicine, Marshall University Huntington, WV; Department of Physiology, University of Arizona, Tucson AZ 85721

## Abstract

**Introduction:** The burden of comorbidities in asthma patients significantly affects management strategies and outcomes. This study aimed to analyze the prevalence and trends of comorbidities among adults with asthma and to investigate their association with persistent asthma.

**Methods:** The study employed data from the Asthma Call-Back Survey (ACBS) to ascertain the prevalence and trends of comorbidities in adults with asthma. Comorbidities were self-reported binary responses. Asthma severity was categorized as intermittent or persistent based on established methodologies to derive asthma severity in the ACBS. Intermittent asthma includes those with current asthma who are well-controlled without being on long-term control medication (LTCM). Persistent asthma includes those on LTCM, regardless of asthma control status, and those not on LTCM whose asthma is not well controlled or is very poorly controlled. Weighted logistic regression controlling for confounders was used to determine the association of comorbidities and asthma severity.

**Results:** Prevalence of comorbidities in adults with asthma were as follows: hypertension (38.4%), major depressive disorder (35.2%), diabetes (17.2%), MI (5.3%), Angina/CHD (6.0%), Stroke (5.0%), and Emphysema/Chronic bronchitis/COPD (19.0%). Prevalence of all comorbidities were higher among adults with persistent asthma compared to intermittent asthma. MI, Angina/CHD, obesity, depression, COPD/emphysema/chronic bronchitis, and hypertension were associated with increased odds of persistent asthma. No association was found between diabetes, stroke, and persistent asthma.

**Conclusion:** Comorbidities are associated with persistent asthma. These findings suggest a need for comprehensive healthcare strategy that address these intertwined health conditions along asthma.

## Introduction

Asthma is a chronic respiratory disorder that affects over 21 million adults in the United States (US) [1] and a substantial proportion of the global population [2], with a wide range of clinical presentations and disease severity [3]. In recent years, there has been a growing recognition of the complex association between asthma and various comorbid conditions [4], which can significantly impact disease management [5], treatment outcomes [6], and overall quality of life [7,8]. Severe asthma, a more critical clinical subset of asthma, affects roughly 10% of the asthma population yet disproportionately shoulders the majority of associated healthcare costs [9]. As per the standards outlined by the European Respiratory Society (ERS) and the American Thoracic Society (ATS), severe asthma necessitates an escalated level of management or persists as uncontrolled despite strenuous therapeutic interventions [10]. Individuals with severe asthma present a broad array of clinical phenotypes and demonstrate diverse treatment responses, underscoring the heterogeneity of this disorder [11].

Comorbid conditions have garnered increased recognition for their potential role in influencing the trajectory of disease progression and the efficacy of treatment measures [12, 13]. Although comorbidities are prevalent among patients with asthma, overall, understanding their prevalence by asthma severity has not been explored within the US population using publicly available data. Understanding the prevalence of these comorbidities in adults with asthma and the association of these comorbidities with asthma severity is crucial for improving clinical care, optimizing therapeutic strategies, and enhancing patient outcomes [14].

Cardiovascular diseases (CVD) like hypertension [15], Myocardial Infarction (MI) [16], coronary heart disease (CHD) [17], and stroke [18] occur in asthmatics and those with severe asthma due to chronic airway inflammation leading to systemic inflammation and increased vulnerability to vascular diseases [19]. Studies have shown that inflammatory biomarkers that are increased in atherogenesis, such as high-sensitivity C-reactive protein (Hs-CRP), interleukin-6 (IL-6), tumor necrosis factor-alpha (TNF-a), interleukin-8 (IL-8), and fibrinogen, are also known to be elevated in asthmatics, ultimately leading to the vascular dysfunction that herald CVD [19,20]. This could in turn exacerbate respiratory symptoms and potentially impact the patient’s overall disease severity [20]. Metabolic disorders such as diabetes [21] and overweight/obesity [22], which are known to increase systemic inflammation and immune dysregulation [23], could influence both the severity of asthma and the individual’s response to treatment [24, 25]. Mental health disorders, particularly major depressive disorder [26,27,28] and poor mental health status [29], amplify symptom perception and complicate effective disease management [28]. Asthma patients experiencing mental health disorders may struggle with medication adherence [27,29], the ability to perform self-care tasks [27], and interpreting the severity of their symptoms [26]. Concomitant respiratory pathologies like Chronic Obstructive Pulmonary Disease (COPD), intensify respiratory impairment and symptom severity [30]. The co-existence of COPD and asthma, often referred to as Asthma-COPD Overlap Syndrome (ACOS), represents a particularly challenging phenotype associated with heightened morbidity and mortality [31,32].

This present study, therefore, aims to:

1. Determine the prevalence of comorbidities among US adults with asthma and their distribution by asthma severity.

2. Investigate the association of comorbidities with asthma severity after controlling for confounders.

## 2. Materials and Methods

### 2.1. Study Population

The present analysis utilized the 2017, 2018, 2019, and 2020 Behavioral Risk Factor Surveillance System (BRFSS) Asthma Call-Back Surveys (ACBS) as the primary data source [33]. The BRFSS is a state-based surveillance system sponsored by the Centers for Disease Control and Prevention (CDC) that collects comprehensive information on chronic diseases, injuries, preventive health practices, health risk behaviors, and healthcare access and utilization, encompassing chronic diseases. The ACBS, conducted two weeks after the BRFSS, is a continuous survey providing annual data on individuals with asthma, including demographics, medical history, asthma risk factors, and medication usage history. The ACBS serves as a vital resource for addressing critical inquiries pertaining to the health status and experiences of individuals with asthma, and it offers data at the state and local levels. Eligibility for ACBS is limited to respondents who self-report a physician diagnosis of asthma in the BRFSS.

To assess the prevalence of asthma in the BRFSS, two questions were employed. An affirmative response to the questions “Have you ever been told by a doctor, nurse, or other healthcare professional that you have asthma?” and “Do you currently have asthma?” were used to indicate lifetime asthma and current asthma status, respectively. Responding affirmatively to the first question was a requirement for participating in the follow-up ACBS. The response rates for ACBS are calculated according to the guidelines set forth by the Council of American Survey and Research Organization (CASRO). In the 2017, 2018, 2019, and 2020 surveys, the ACBS response rates for participating states, colonies, and Washington, DC, were 45%, 41.6%, 49.4%, and 49.7%, respectively [33].

The study population consisted of 48,104 adults with asthma who participated in the ACBS across 31 states and Puerto Rico. Sample survey weights were adjusted as per the recommendations of the ACBS to account for survey design and ensure representativeness. The ACBS received approval from the Institutional Review Boards of the respective states and the Ethics Review Board of the Asthma and Community Health Branch of the National Center for Environmental Health. Informed consent was obtained from all participants. Since the ACBS dataset is publicly available, this study is exempt from full institutional review board review. Detailed information on the data, sampling methods, and analytical guidelines can be found on the ACBS website at https://www.cdc.gov/brfss/acbs/index.htm [33].

### 2.2. Asthma severity ascertainment

The determination of asthma severity in this study followed established methodologies consistent with previous publications by the CDC that utilized data from the ACBS to derive population-level asthma severity estimates [34, 35]. First, respondents were classified into three categories of asthma control: well-controlled, not well-controlled, and very poorly controlled. This classification was based on three measures of impairment reported by the respondents: daytime symptoms experienced in the past 30 days, nighttime symptoms in the past 30 days, and use of short-acting beta 2-agonists (SABA) for symptom control during the previous 3 months (excluding prevention of exercise-induced bronchospasm). The specific criteria and definitions used for this classification are provided in Supplementary Table 1 [34]. This methodology used a modified version of the 2007 National Asthma Education and Prevention Program (NAEPP) guidelines, as some required measures for assessing current impairment and future risk assessment were not available in the ACBS dataset [36]. Secondly, the use of long-term asthma control medications, including inhaled and systemic corticosteroids, long-acting beta 2-agonists, leukotriene receptor antagonists, methylxanthines, and immunomodulators, was determined based on the reported medication history. This information was categorized as a binary variable (yes or no) to indicate the presence or absence of long-term control medication usage [34, 35]. Using the assessment of asthma control status and long-term control medication usage, asthma severity was then classified into two categories: intermittent and persistent asthma. Intermittent asthma includes individuals with current asthma who were well controlled without the need for long-term control medications. Persistent asthma includes individuals who require long-term control medications, irrespective of their asthma control status, as well as those who did not utilize long-term control medications but had poorly controlled or very poorly controlled asthma [34]. Additional details regarding the classification criteria can be found in Supplementary Table 2.

### 2.3. Assessment of Comorbidities

Comorbidities were assessed in BRFSS via the chronic conditions questionnaire module that collected self-reported information pertaining to individuals’ medical history and overall health status. To ascertain the presence of comorbidities, participants were asked the following question: "Has a doctor, nurse, or other health professional ever informed you that you have any of the following conditions?" Comorbidities assessed through this method included Angina/Coronary Heart Disease, Stroke, Diabetes, Myocardial Infarction, Major Depressive disorder, Chronic Obstructive Pulmonary Disease (COPD), Emphysema, Chronic Bronchitis, and Hypertension. Responses were recorded as binary variables (Yes vs No). Participants who responded as "Don’t know" or who refused to answer the question were coded as missing data for the variable. For hypertension, females who reported hypertension exclusively during pregnancy and adults who were informed they had borderline high or pre-hypertensive disorder were also coded as missing. Comorbid overweight and obesity were assessed based on body mass index (BMI), calculated from self-reported height and weight measurements. BMI was categorized as follows: Underweight (BMI < 18.5 kg/m^2), Normal weight (BMI ≥ 18.5 to 24.9 kg/m^2), Overweight (BMI ≥ 25 to 29.9 kg/m^2), and Obesity (BMI ≥ 30 kg/m^2). Underweight adults were recorded as having missing values for BMI. To assess poor mental health status, participants were asked the following survey question: "Now thinking about your mental health, which includes stress, depression, and problems with emotions, how many days during the past 30 days was your mental health not good?". The responses were categorized as "zero days when mental health was not good," "1-14 days when mental health was not good," and "≥ 15 days when mental health was not good." The CDC Mental Health Days questionnaire is a validated and consistently used measure for assessing mental health status and Health Related Quality of life in adults [37].

### 2.4. Confounders

Several potential confounding variables were considered based on a prior study [38], including age (categorized as 18–34 years, 35–44 years, 45–54 years, 55–64 years, or ≥65 years), gender (male or female), race (white non-Hispanic, Black non-Hispanic, Hispanic, or multiracial/other non-Hispanic), household smoke exposure (yes or no), smoking status (never smokers, former smokers, and current smokers), household income (≥$25,000 or <$25,000), the presence of mold in the home (yes or no), educational status (college graduate/higher, some college, high school or less), health care coverage in past 12 months (none or yes), physical activity or exercise during the past 30 days other than regular job (yes or no), having an asthma management plan (yes or no). Heavy drinking was defined in the BRFSS as the consumption of more than 14 drinks per week for adult men or more than 7 drinks per week for adult women. This variable was treated as a binary yes or no variable.

### 2.5. Statistical Analysis

The analysis of the Asthma Call-Back Survey (ACBS) data incorporated appropriate statistical techniques to account for the complex survey design and ensure the representativeness of the U.S. adult population with asthma. The survey employed a stratified, multistage complex sampling design, and analytical guidelines recommended the use of PROC SURVEYFREQ and PROC SURVEYLOGISTIC procedures. Weighting procedures were applied to address the sampling strata, cluster, and primary sampling unit (PSU) in order to provide accurate and reliable estimates. All reported proportions were expressed as weighted percentages. The prevalence of comorbidities was assessed within each ACBS cycle and in the overall sample. Prevalence ratios were calculated to compare the prevalence of comorbidities between specific years (e.g., 2019 vs. 2008) and evaluate trends. Trends in comorbidities were reported as decreasing (prevalence ratio < 1, p for trend < 0.05), increasing (prevalence ratio > 1, p for trend < 0.05), or stable (p for trend ≥ 0.05) [39].

To assess the association of comorbidities with asthma severity, weighted multi-logistic regression models were employed. Each comorbidity was independently regressed on the outcome while controlling for confounders. The results of the logistic regression models were reported as odds ratios (O.R.s) along with their corresponding 95% confidence intervals (C.I.s). Multicollinearity among the independent variables was assessed through tolerance estimates and Variance Inflation Factor (VIF) calculations. Multicollinearity was considered absent if no tolerance estimate was below 0.1 and no VIF exceeded 10 [40]. Statistical hypotheses were evaluated using a predetermined level of significance set at a two-sided p-value < 0.05 or non-overlapping 95% confidence intervals. Data analysis was conducted using the SAS 9.4 statistical software (SAS Institute, Cary, NC, USA).

## Results

### Prevalence and trends of comorbidities among adults with asthma

The proportion of respondents with comorbidities were as follows: MI (5.3%), Angina/CHD (6.0%), Stroke (5.0%), Diabetes (17.2%), Major depressive disorder (35.2%), Emphysema (5%), Chronic bronchitis (19.9%), COPD (12.5%), hypertension (38.4%), 1-14 poor mental health days (32.2%) and ≥15 poor mental health days (20.7%). The prevalence of having either COPD or emphysema or chronic bronchitis was 19.0% (Table 1, Figure 1). Trends of comorbidities were stable across ACBS cycles (p for trend ≥ 0.05) (Table 2, Figure 4).

**Figure 1:**
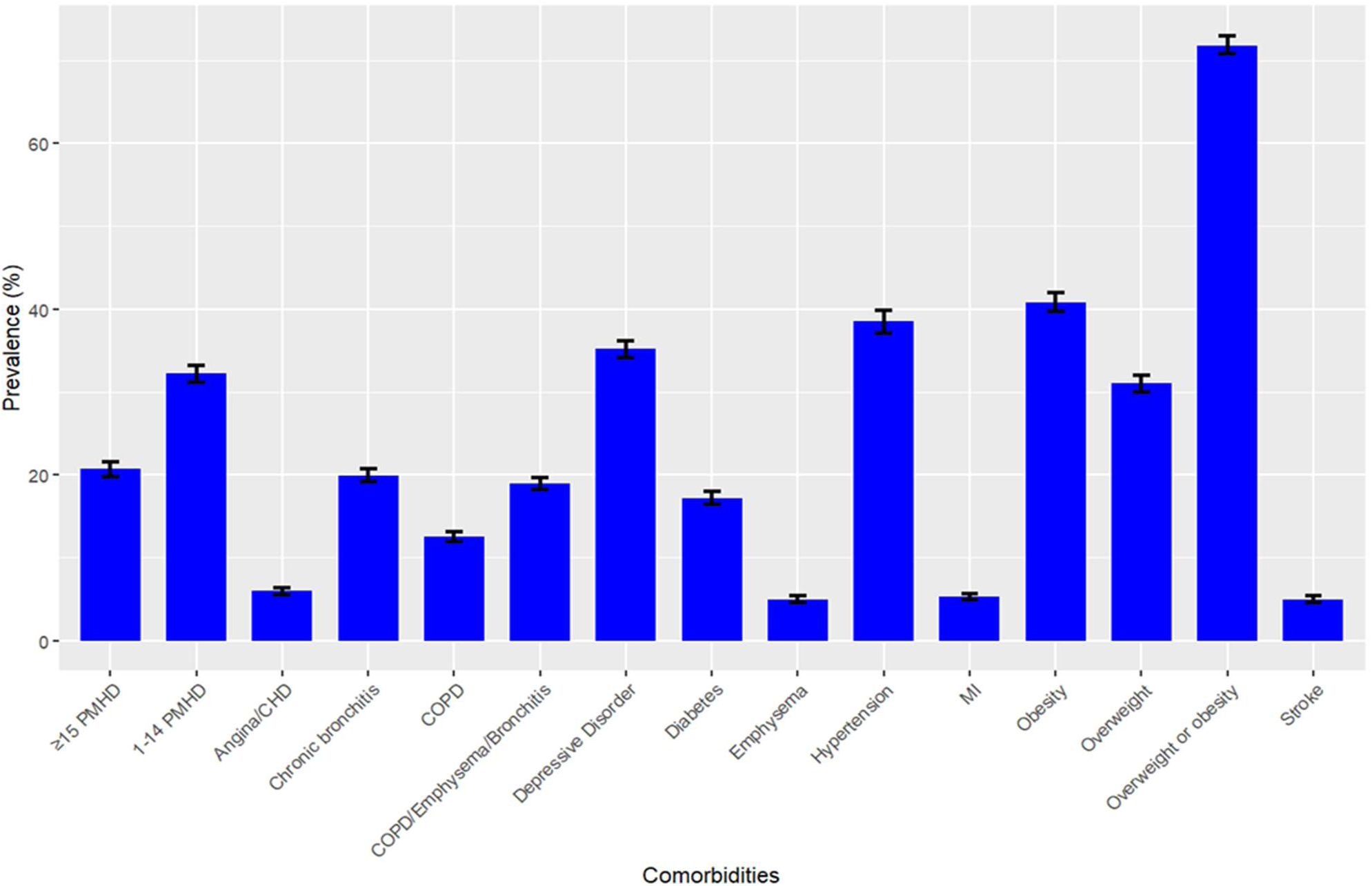
Prevalence of Comorbidities in US Adults with Asthma, 2017-2021 Asthma Call-Back Survey.

**Figure 2:**
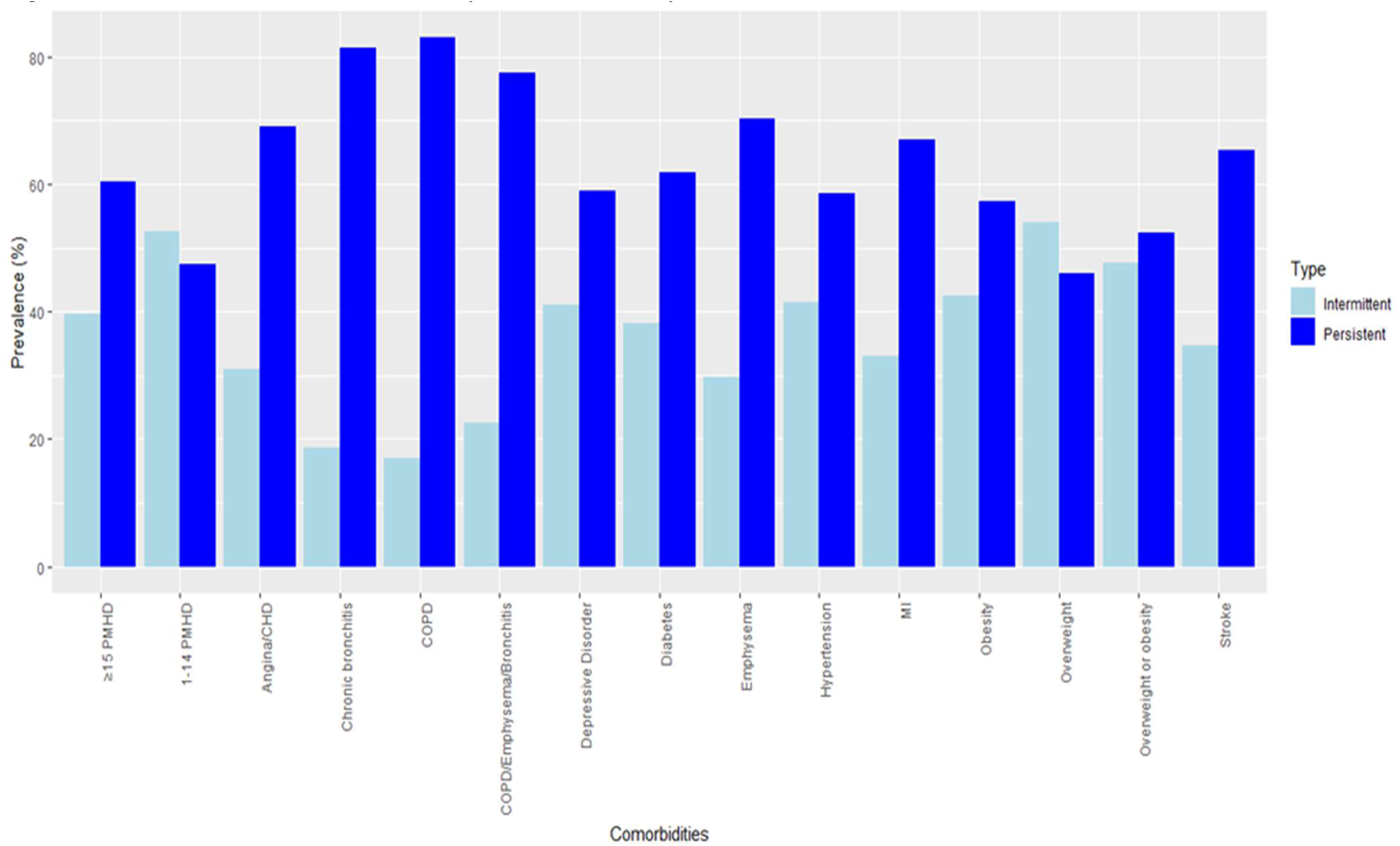
Prevalence of Comorbidities by Asthma Severity Status (Intermittent or Persistent)

**Figure 3:**
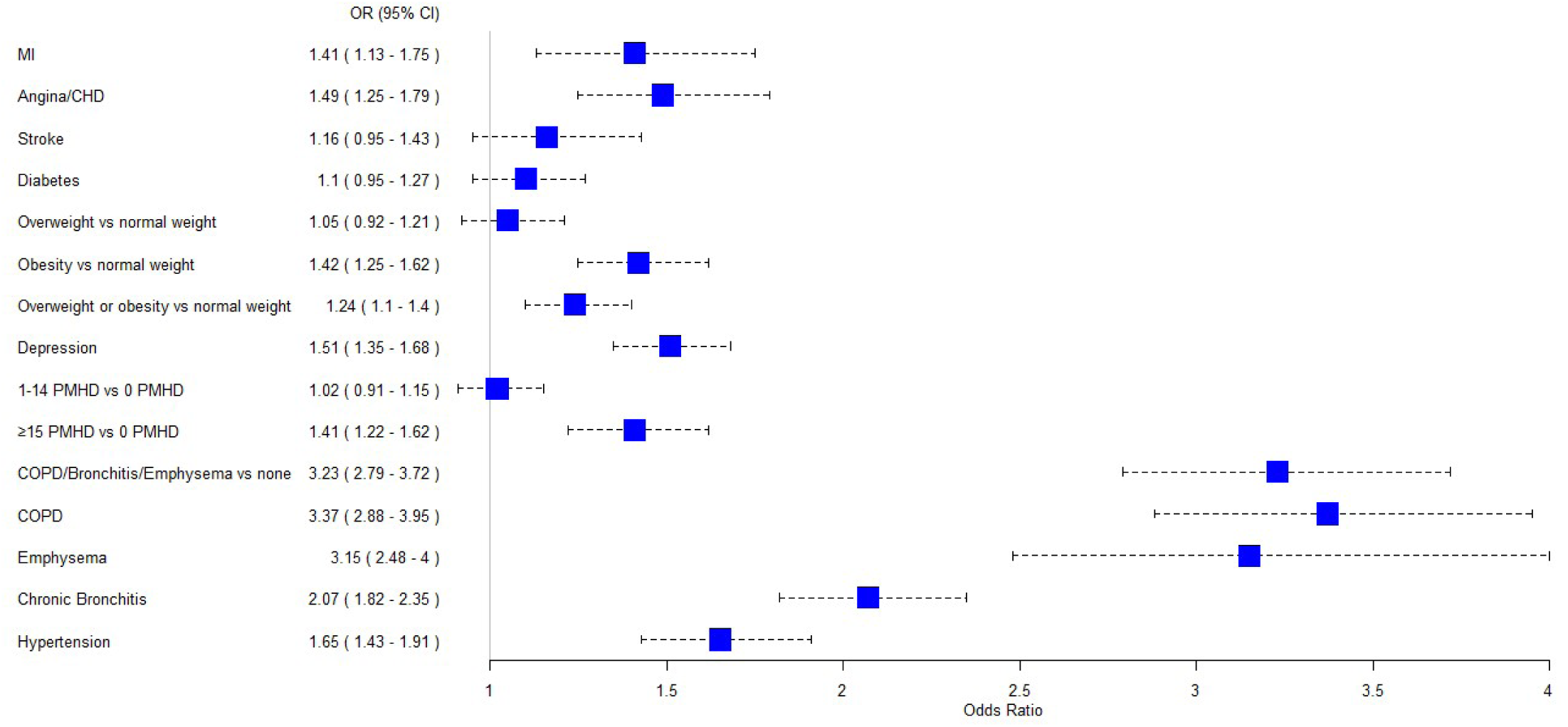
Weighted Multivariable Logistic Regression Analysis of the Association between Comorbidities and Asthma Severity MI, Myocardial Infarction; COPD, Chronic Obstructive Pulmonary Disease; PMHD, Poor Mental Health Days; CHD, Coronary Heart Disease. Each comorbidity was controlled for age, gender, race, household smoke exposure, smoking status, household income, presence of mold in the home, educational level, health care coverage in past 12 months, physical activity/exercise during the past 30 days other than regular job, having asthma management plan, and heavy drinking.

**Figure 4:**
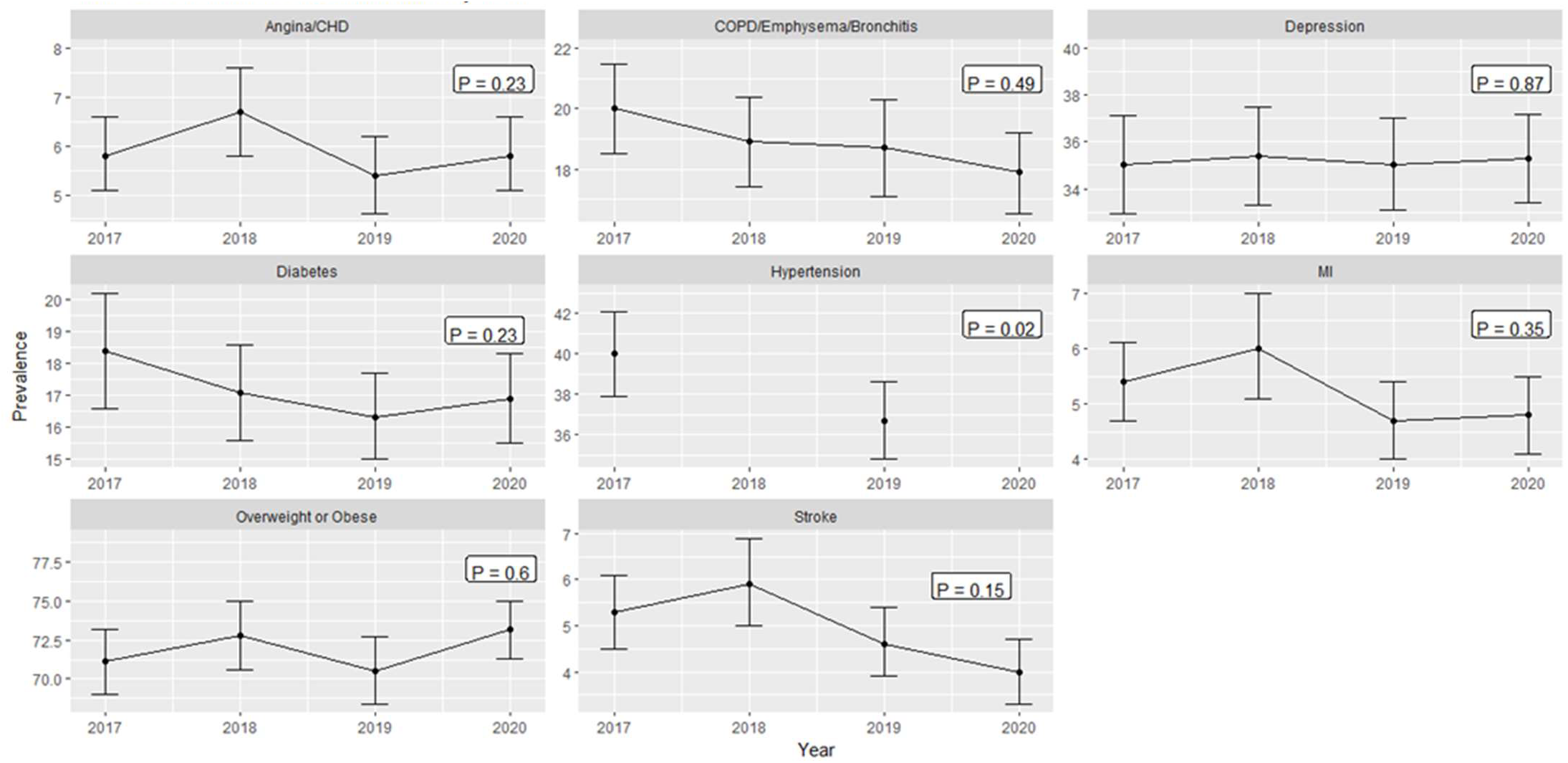
Trends in Prevalence of Comorbidities by Year

**Table 1:** Descriptive Statistics of Risk Factors and Comorbidities among US Adults with Asthma Stratified by Asthma Severity.

| Characteristics | Survey Respondents |  | Intermittent Asthma |  | Persistent Asthma |  | P - value |
| --- | --- | --- | --- | --- | --- | --- | --- |
|  | No.<sup>a</sup> | %<sup>b</sup> | No.<sup>a</sup> | %<sup>b</sup> | No.<sup>a</sup> | %<sup>b</sup> |  |
| **Total** | 48104 |  | 21123 | 50.4 | 26981 | 49.6 | 0.47 |
| **Sex** |  |  |  |  |  |  | <.0001 |
| Female | 30651 | 59.4 | 12045 | 44.3 | 18606 | 55.7 |  |
| Male | 17453 | 40.6 | 9078 | 59.3 | 8375 | 40.7 |  |
| **Age group** |  |  |  |  |  |  | <0.0001 |
| 18-34 years | 7864 | 33.1 | 4837 | 63.9 | 3027 | 36.1 |  |
| 35-44 years | 5168 | 14.9 | 2695 | 53.3 | 2473 | 46.7 |  |
| 45-54 years | 7121 | 16.5 | 2990 | 43.4 | 4131 | 56.6 |  |
| 55-64 years | 11144 | 16.7 | 4138 | 40 | 7006 | 60 |  |
| 65+ years | 16807 | 18.7 | 6463 | 39.4 | 10344 | 60.6 |  |
| **Race/ethnicity** |  |  |  |  |  |  | <0.0001 |
| White, non-Hispanic | 38031 | 66.9 | 16497 | 48.6 | 21534 | 51.4 |  |
| Black, non-Hispanic | 2597 | 10.4 | 1113 | 51.7 | 1484 | 48.3 |  |
| Hispanic | 3824 | 15.4 | 1857 | 54.7 | 1967 | 45.3 |  |
| Multiple/other race, non-Hispanic | 3652 | 7.4 | 1656 | 56 | 1996 | 44 |  |
| **Household income** |  |  |  |  |  |  | <.0001 |
| ≥\$25,000 | 29077 | 70.2 | 14210 | 54.5 | 14867 | 45.5 | |
| <\$25,000 | 12850 | 29.8 | 4258 | 40.8 | 8592 | 59.2 | |
| **Education** |  |  |  |  |  |  | <.0001 |
| College graduate/higher | 19185 | 29.8 | 9363 | 55.7 | 9822 | 44.3 |  |
| Some college | 14859 | 35 | 6270 | 51 | 8589 | 49 |  |
| High school or less | 13997 | 35.2 | 5473 | 45.3 | 8524 | 54.7 |  |
| **Health care coverage in past 12 months** |  |  |  |  |  |  | 0.0033 |
| None | 2873 | 8.2 | 1485 | 56.4 | 1388 | 43.6 |  |
| Yes | 45112 | 91.8 | 19570 | 49.8 | 25542 | 50.2 |  |
| **Smoking status** |  |  |  |  |  |  | <.0001 |
| Never smoker | 26437 | 58.9 | 12568 | 54.7 | 13869 | 45.3 |  |
| Former smoker | 14422 | 25.3 | 6027 | 46.6 | 8395 | 53.4 |  |
| Current smoker | 7028 | 15.7 | 2429 | 40.6 | 4599 | 59.4 |  |
| **Household smoke exposure** |  |  |  |  |  |  | <.0001 |
| No | 42529 | 87.1 | 19199 | 51.7 | 23330 | 48.3 |  |
| Yes | 5538 | 12.9 | 1906 | 41.6 | 3632 | 58.4 |  |
| **Mold in the home** |  |  |  |  |  |  | <.0001 |
| No | 43265 | 90.4 | 19484 | 51.4 | 23781 | 48.6 |  |
| Yes | 4613 | 9.6 | 1564 | 41.5 | 3049 | 58.5 |  |
| **Physical activity** |  |  |  |  |  |  | <.0001 |
| No | 13826 | 26.4 | 4962 | 40.6 | 8864 | 59.4 |  |
| Yes | 34210 | 73.6 | 16138 | 53.9 | 18072 | 46.1 |  |
| **Heavy drinker** |  |  |  |  |  |  | 0.102 |
| No | 44749 | 93.8 | 19559 | 50.1 | 25190 | 49.9 |  |
| Yes | 2699 | 6.2 | 1276 | 53.9 | 1423 | 46.1 |  |
| **Have an asthma management plan** |  |  |  |  |  |  | <.0001 |
| No | 33627 | 71 | 15798 | 53.3 | 17829 | 46.7 |  |
| Yes | 13262 | 29 | 4616 | 41.4 | 8646 | 58.6 |  |
| **Comorbidities** |  |  |  |  |  |  |  |
| **Myocardial Infarction** |  |  |  |  |  |  | <.0001 |
| No | 44064 | 94.7 | 19874 | 51.4 | 24190 | 48.6 |  |
| Yes | 3755 | 5.3 | 1168 | 33.1 | 2587 | 66.9 |  |
| **Angina/Coronary Heart Disease** |  |  |  |  |  |  | <.0001 |
| No | 43386 | 94 | 19671 | 51.7 | 23715 | 48.3 |  |
| Yes | 4211 | 6 | 1300 | 30.9 | 2911 | 69.1 |  |
| **Stroke** |  |  |  |  |  |  | <.0001 |
| No | 44881 | 95 | 20080 | 51.3 | 24801 | 48.7 |  |
| Yes | 3018 | 5 | 974 | 34.7 | 2044 | 65.3 |  |
| **Diabetes** |  |  |  |  |  |  | <.0001 |
| No | 37345 | 82.8 | 17185 | 53 | 20160 | 47 |  |
| Yes | 10195 | 17.2 | 3690 | 38.2 | 6505 | 61.8 |  |
| **Body Mass Index Status** |  |  |  |  |  |  | <.0001 |
| Normal weight | 11319 | 28.2 | 5642 | 57.2 | 5677 | 42.8 |  |
| Overweight | 14290 | 31 | 6746 | 54.1 | 7544 | 45.9 |  |
| Obesity | 19458 | 40.8 | 7513 | 42.6 | 11945 | 57.4 |  |
| **Major Depressive Disorder** |  |  |  |  |  |  | <.0001 |
| No | 30718 | 64.8 | 14790 | 55.4 | 15928 | 44.6 |  |
| Yes | 17117 | 35.2 | 6215 | 41.0 | 10902 | 59.0 |  |
| **COPD or Emphysema or Bronchitis** |  |  |  |  |  |  | <.0001 |
| No | 35958 | 81 | 18511 | 57.1 | 17447 | 42.9 |  |
| Yes | 11712 | 19 | 2503 | 22.6 | 9209 | 77.4 |  |
| **Emphysema** |  |  |  |  |  |  | <.0001 |
| No | 44269 | 95 | 20354 | 52.2 | 23915 | 47.8 |  |
| Yes | 3251 | 5 | 550 | 17 | 2701 | 83 |  |
| **Chronic bronchitis** |  |  |  |  |  |  | <.0001 |
| No | 35841 | 80.1 | 17748 | 55.7 | 18093 | 44.3 |  |
| Yes | 11471 | 19.9 | 3056 | 29.7 | 8415 | 70.3 |  |
| **COPD** |  |  |  |  |  |  | <.0001 |
| No | 38824 | 87.5 | 19311 | 55.2 | 19513 | 44.8 |  |
| Yes | 8619 | 12.5 | 1569 | 18.6 | 7050 | 81.4 |  |
| **Poor mental health status** |  |  |  |  |  |  | <.0001 |
| 0 poor mental health days | 24769 | 47.1 | 11637 | 54.1 | 13132 | 45.9 |  |
| 1-14 poor mental health days | 13764 | 32.2 | 6281 | 52.5 | 7483 | 47.5 |  |
| ≥15 poor mental health days | 8842 | 20.7 | 2944 | 39.7 | 5898 | 60.3 |  |
| **Hypertension <sup>c</sup>** |  |  |  |  |  |  | <.0001 |
| No | 12120 | 61.6 | 5995 | 56.1 | 6125 | 43.9 |  |
| Yes | 11675 | 38.4 | 4304 | 41.4 | 7371 | 58.6 |  |

CI, confidence interval; COPD, Chronic Obstructive Pulmonary Disease
<sup>a</sup> Unweighted pooled sample size, 2017–2020. Due to item non-response, individual characteristic categories may not sum to total.
<sup>b</sup> Weighted prevalence and 95% confidence interval.
<sup>c</sup> Variable available in only 2018 and 2020 ACBS Dataset

**Table 2:**
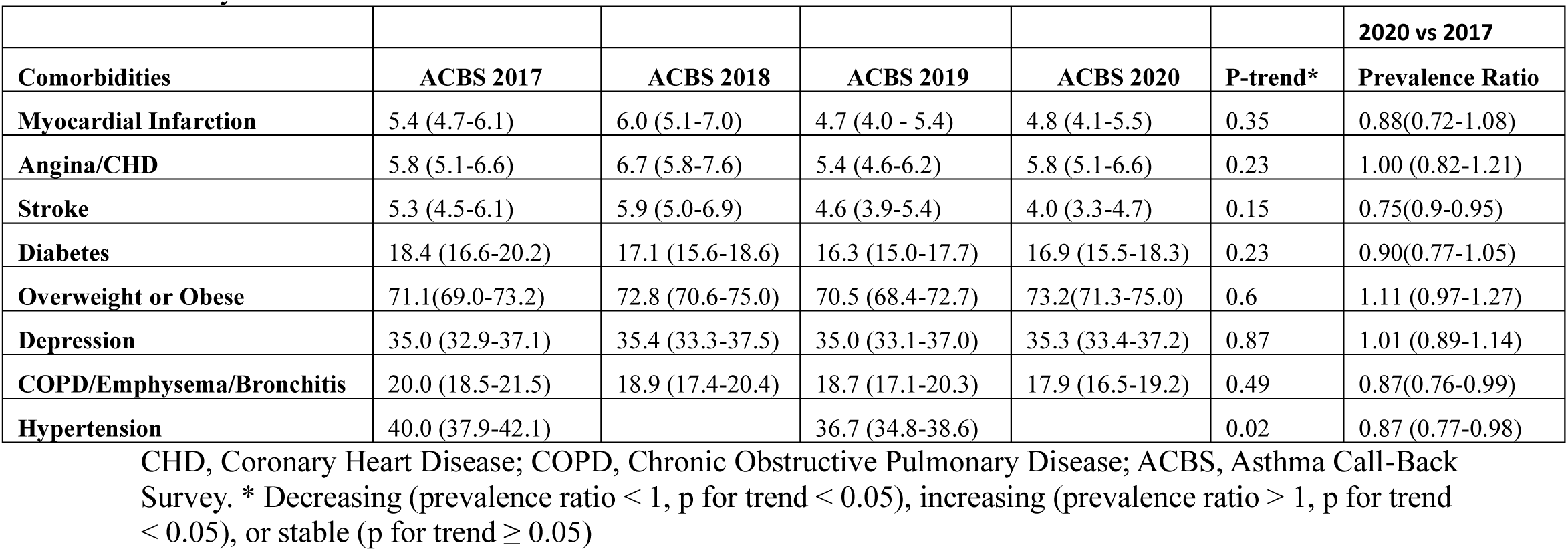
Trend by Cycle Year Prevalence of Comorbidities in US Adults with Asthma, 2017-2021 Asthma Call-Back Survey.

|  |  |  |  |  |  | **2020 vs 2017** |
| --- | --- | --- | --- | --- | --- | --- |
| **Comorbidities** | **ACBS 2017** | **ACBS 2018** | **ACBS 2019** | **ACBS 2020** | **P-trend\*** | **Prevalence Ratio** |
| **Myocardial Infarction** | 5.4 (4.7-6.1) | 6.0 (5.1-7.0) | 4.7 (4.0 - 5.4) | 4.8 (4.1-5.5) | 0.35 | 0.88(0.72-1.08) |
| **Angina/CHD** | 5.8 (5.1-6.6) | 6.7 (5.8-7.6) | 5.4 (4.6-6.2) | 5.8 (5.1-6.6) | 0.23 | 1.00 (0.82-1.21) |
| **Stroke** | 5.3 (4.5-6.1) | 5.9 (5.0-6.9) | 4.6 (3.9-5.4) | 4.0 (3.3-4.7) | 0.15 | 0.75(0.9-0.95) |
| **Diabetes** | 18.4 (16.6-20.2) | 17.1 (15.6-18.6) | 16.3 (15.0-17.7) | 16.9 (15.5-18.3) | 0.23 | 0.90(0.77-1.05) |
| **Overweight or Obese** | 71.1(69.0-73.2) | 72.8 (70.6-75.0) | 70.5 (68.4-72.7) | 73.2(71.3-75.0) | 0.6 | 1.11 (0.97-1.27) |
| **Depression** | 35.0 (32.9-37.1) | 35.4 (33.3-37.5) | 35.0 (33.1-37.0) | 35.3 (33.4-37.2) | 0.87 | 1.01 (0.89-1.14) |
| **COPD/Emphysema/Bronchitis** | 20.0 (18.5-21.5) | 18.9 (17.4-20.4) | 18.7 (17.1-20.3) | 17.9 (16.5-19.2) | 0.49 | 0.87(0.76-0.99) |
| **Hypertension** | 40.0 (37.9-42.1) |  | 36.7 (34.8-38.6) |  | 0.02 | 0.87 (0.77-0.98) |

CHD, Coronary Heart Disease; COPD, Chronic Obstructive Pulmonary Disease; ACBS, Asthma Call-Back Survey. \* Decreasing (prevalence ratio < 1, p for trend < 0.05), increasing (prevalence ratio > 1, p for trend < 0.05), or stable (p for trend ≥ 0.05)

### Prevalence of comorbidities by asthma severity

Among adults with comorbidities, prevalence of persistent asthma was higher in those with obesity (57.4%), Diabetes (65.3%), Stroke (65.3%), Angina/CHD (69.1%), MI (66.9%), major depressive disorder (59.0%), emphysema (83%), chronic bronchitis (70.3%), COPD (81.45), COPD or Emphysema or Bronchitis (77.4%), hypertension (58.6%), and ≥15 poor mental health days (60.3%) (Table 1, Figure 2).

### Association of comorbidities with asthma severity

MI was associated with persistent asthma (Adjusted Odds Ratio, aOR=1.41 (95% Confidence Interval, CI=1.13-1.75) (Figure 3). Endorsing Angina/CHD compared with none was associated with 49% increased odds of persistent asthma (aOR = 1.49, 95% CI = 1.25–1.79). Obese adults, compared to normal-weight adults, were more likely to have persistent asthma (aOR = 1.42, 95% CI = 1.25–1.62). A history of depression and ≥15 PMHD were associated with increased odds of persistent asthma, respectively (aOR = 1.51, 95% CI = 1.35–1.68; aOR = 1.41, 95% CI = 1.22– 1.62). Compared with those without COPD, those with COPD had 237% increased odds of persistent asthma (aOR = 3.37, 95% CI = 2.88–3.95). Emphysema and chronic bronchitis were associated with 215% and 107% increased odds of persistent asthma, respectively (aOR = 3.15, 95% CI = 2.48–4.0; aOR = 2.07, 95% CI = 1.82–2.35). Hypertension was associated with persistent asthma (aOR = 1.65, 95% CI = 1.43–1.91). We did not find an association between diabetes, stroke, and persistent asthma (p>0.05).

## Discussion

The present study aimed to evaluate the prevalence of comorbidities within the adult asthma population in the United States, with a particular emphasis on delineating the prevalence in individuals with intermittent and persistent asthma, as well as examining the association of these comorbidities with persistent asthma. We observed an increased prevalence of comorbidities in adults with asthma, corroborating the findings of Tsai et al. (2020), who observed a prevalence range of 2.0%-12% for cardiovascular and cerebrovascular comorbidities such as hypertension, myocardial infarction, angina, and stroke among adults aged 18 and above [41]. Conversely, higher prevalence rates have been reported in other studies, attributed to heterogeneity in the study population and settings, including the inclusion of institutionalized clinical patients [42, 43,44]. In addition, the prevalence of poor mental health status, depression, obesity, chronic obstructive pulmonary disease (COPD), and emphysema was similar to what has been reported in another study [4].

The prevalence of all comorbidities was higher in adults with persistent asthma compared to those with intermittent asthma. An association between COPD and persistent asthma was noted, which is substantiated by earlier studies that indicated an augmented risk of chronic bronchitis [45,46] and COPD [47] in asthma patients compared to non-asthma counterparts. The underlying mechanistic interplay between COPD and asthma remains a subject of scholarly debate; however, theories posit that obstructive lung disease may have a shared genetic pathway, with bronchial hyperresponsiveness playing a pivotal role in both asthma and COPD [48]. Additionally, heightened inflammation is observed among adults with persistent asthma, and environmental factors such as smoking may modulate this pathway [48].

We found that MI, Angina/CHD, and hypertension were associated with persistent asthma. A multicenter cohort study showed that individuals with persistent asthma had the lowest unadjusted cardiovascular disease (CVD)-free survival rate compared to their intermittent asthmatic and non-asthmatic counterparts [49]. While no elevated CVD risk was discerned among intermittent asthmatics vs non-asthmatics, a higher risk of CVD events was detected among persistent asthmatics in comparison to non-asthmatics after accounting for confounding variables [49]. This alludes to a potential underlying pathophysiological nexus between cardiovascular disease and asthma. Severe asthma may foster the generation of procoagulant factors encompassing thrombin, fibrin, and soluble tissue factors, further potentiating the prothrombotic state via impairment of the protein C anticoagulant system and fibrinolysis [50]. Additionally, chronic inflammatory states as seen in severe asthma may contribute to the compromise of vascular endothelial lining integrity [19].

A significant association between depression, poor mental health status, and obesity with persistent asthma persisted after adjusting for confounders. Cross-sectional studies have consistently documented an increased likelihood of reporting asthma in individuals with depression and poor mental health status [28, 29, 51]. A national cohort study conducted in Korea highlighted a bidirectional relationship between asthma and depression, suggesting a deeper underlying mechanism [28]. It is postulated that the dysregulation of the pro-anti-inflammatory balance [52] and cytokine imbalance [53] prevalent in depression and other psychopathologies are associated with elevated systemic levels of inflammatory mediators such as interleukin-4, IL-6, and tumor necrosis factor-alpha, which are also implicated in the pathogenesis of asthma [53]. Historically, the relationship between obesity and asthma severity was perceived as unidirectional, where individuals with severe asthma, became less active and deconditioned, gain weight, a situation further aggravated by increased usage of oral corticosteroids [54]. However, recent evidence suggests that adult body mass index may play a causal role in the risk of asthma, with the effect of asthma on body mass index being trivial, if at all present [55]. This new understanding proposes that obesity undermines optimal health by inciting adipose tissue to release inflammatory mediators due to an excess of macronutrients and by decreasing the production of adiponectin, which predisposes individuals to a proinflammatory state [56].

Our study identified an absence of a significant association between diabetes, stroke, and persistent asthma. The treatment of severe asthma often involves repeated or sustained use of oral glucocorticoids, which are purported to increase the risk of type 2 diabetes [57], though this correlation is not consistently observed across all studies [58, 59]. Furthermore, a considerable prevalence of asthma has been detected among patients with type 2 diabetes, as confirmed in a large-scale Danish twin study [60]. Taken together, these studies indicate that the causal link between diabetes and asthma could be bidirectional. The relationship between asthma and stroke risk has similarly been scrutinized. Although a meta-analysis concluded that asthma could significantly increase the risk of stroke and its impact was not consistent in different populations, there are no studies that evaluated the risk of stroke outcome by asthma severity status [61]. The lack of a clear association could be potentially ascribed to multiple underlying mechanisms. Given the heterogeneity of both asthma and diabetes as diseases, it is crucial to recognize distinct asthma endotypes, each typified by a variety of pathological processes [62]. It is also plausible that the association of stroke and diabetes with persistent asthma could be obscured by confounding factors such as concurrent cardiovascular disease, other comorbidities, and residual confounders. Future longitudinal studies are warranted to further illuminate the potential links between these comorbidities and the severity of asthma.

This study’s principal strength lies in the employment of a sizable sample of adults presently diagnosed with asthma, focusing on the assessment of asthma severity within states contributing to the Asthma Call-Back Survey (ACBS). The ACBS holds a distinctive position as the singular survey supplying the bulk of indicators necessary for defining asthma severity and control, consistent with the National Asthma Education and Prevention Program (NAEPP, EPR-3) guidelines. Consequently, the ACBS serves as an essential instrument for evaluating asthma severity and control at a population scale.

In interpreting the results disclosed in this research, it is important to acknowledge certain study limitations. A significant limitation stems from the constraints inherent in the available ACBS indicators utilized for the categorization of asthma severity. The questionnaire content of the ACBS did not permit the integration of all elements stipulated by the NAEPP guidelines, such as activity limitation, pulmonary function measures, and asthma exacerbations requiring oral corticosteroids. This factor could potentially contribute to an underestimation of persistent asthma prevalence. Despite this, the asthma severity classification applied in this analysis concurs with the definitions employed in other population-based estimates [34,35]. Additionally, the range of assessed comorbidities was also delimited by the ACBS. For instance, gastroesophageal reflux disorder, vocal cord dysfunction, obstructive sleep apnea, rhinitis, and atopy, all acknowledged comorbidities linked to asthma [63], were not evaluated. Moreover, each analytical model evaluated a solitary predictor (comorbidity), with adjustments made for confounding variables, presuming a unique model where a single comorbidity serves as an exposure. However, it is pertinent to consider that comorbidities could interact with each other, and their multiplicity could have diverse interactions or associations with asthma severity. Such potential interactions among comorbidities were not investigated in our study. Given the observational and cross-sectional nature of this study, recall bias is unavoidable and a causal relationship between these comorbidities and asthma severity cannot be established. Finally, the generalizability of our findings is limited to adults with current asthma residing in states that participated in the ACBS and does not extend to institutionalized patients.

In conclusion, the prevalence of comorbidities amongst the US adult population diagnosed with asthma were higher in adults with persistent asthma compared with intermittent asthma, and most comorbid conditions were associated with persistent asthma. The identified associations between various comorbidities and persistent asthma underline the complex, multidimensional nature of asthma, pointing towards the necessity for a comprehensive, integrated approach to patient management. This evidence should stimulate healthcare providers to anticipate these comorbidities and adjust their approach to better manage individuals with asthma.

## Data Availability

All data produced are available online at https://www.cdc.gov/brfss/acbs/index.htm

## Institutional Review Board Statement

Not applicable.

## Informed Consent Statement

Not applicable.

## Data Availability Statement

The data used to generate the findings of this study are publicly available in the CDC Asthma Call-Back Survey Website available at: https://www.cdc.gov/brfss/acbs/index.htm (accessed on 1 June 2023). The Asthma Call-back Survey (ACBS) is a product of CDC’s National Asthma Control Program (NACP).

## Supplementary tables

**Supplementary table 1:**
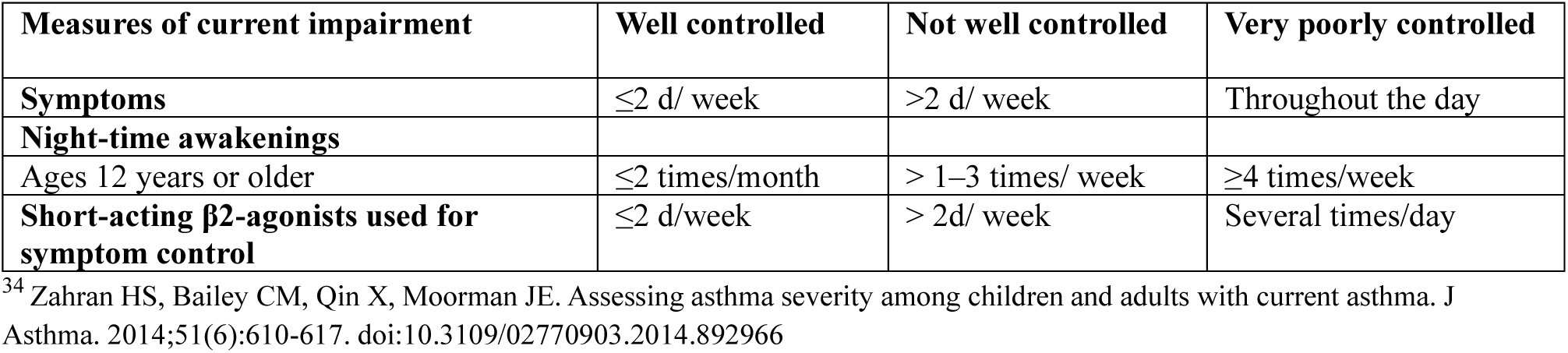
Classification of asthma control in adults modified from the National Asthma Education and Prevention Program Expert Panel Report 3 Guidelines. ^34^.

**Supplementary Table 2:**
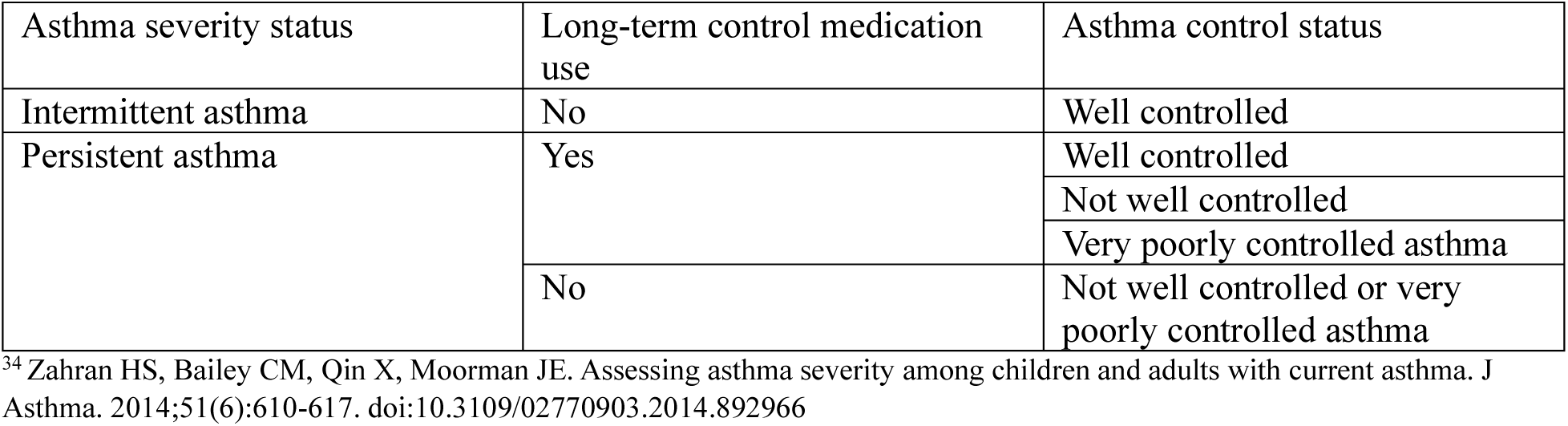
Classification of asthma severity for research and population-based estimates from the National Asthma Education and Prevention Expert Panel Report 3 guidelines ^34^.

