## Supplementary tables for "Prevalence of Comorbidities among United States Adults with asthma and Their Association with Asthma Severity"

Supplementary table 1: Classification of asthma control in adults modified from the National Asthma Education and Prevention Program Expert Panel Report 3 Guidelines. ^34^

| **Measures of current impairment** | **Well controlled** | **Not well controlled** | **Very poorly controlled** |
| --- | --- | --- | --- |
| **Symptoms** | ≤2 d/ week | >2 d/ week | Throughout the day |
| **Night-time awakenings** |  |  |  |
| Ages 12 years or older | ≤2 times/month | > 1–3 times/ week | ≥4 times/week |
| **Short-acting β2-agonists used for symptom control** | ≤2 d/week | > 2d/ week | Several times/day |

^34^ Zahran HS, Bailey CM, Qin X, Moorman JE. Assessing asthma severity among children and adults with current asthma. J Asthma. 2014;51(6):610-617. doi:10.3109/02770903.2014.892966

Supplementary Table 2: Classification of asthma severity for research and population-based estimates from the National Asthma Education and Prevention Expert Panel Report 3 guidelines ^34^

| Asthma severity status | Long-term control medication use | Asthma control status |
| --- | --- | --- |
| Intermittent asthma | No | Well controlled |
| Persistent asthma | Yes | Well controlled |
|  |  | Not well controlled |
|  |  | Very poorly controlled asthma |
|  | No | Not well controlled or very poorly controlled asthma |

^34^ Zahran HS, Bailey CM, Qin X, Moorman JE. Assessing asthma severity among children and adults with current asthma. J Asthma. 2014;51(6):610-617. doi:10.3109/02770903.2014.892966
